# Mathematical modelling to estimate the impact of maternal and perinatal healthcare services and interventions on health in sub-Saharan Africa: A scoping review

**DOI:** 10.1101/2023.12.16.23300088

**Authors:** Joseph H Collins, Valentina Cambiano, Andrew N. Phillips, Tim Colbourn

## Abstract

Mathematical modelling is a commonly utilised tool to predict the impact of policy on health outcomes globally. Given the persistently high levels of maternal and perinatal morbidity and mortality in sub-Saharan Africa, mathematical modelling is a potentially valuable tool to guide strategic planning for health and improve outcomes. The aim of this scoping review was to explore how modelling has been used to evaluate the delivery of maternal and/or perinatal healthcare interventions or services and predict their impact on health-related outcomes in the region. A search across three databases was conducted in November 2023 which returned 8660 potentially relevant studies, from which 60 were included in the final review. Characteristics of these studies, the interventions which were evaluated, the models utilised, and the analyses conducted were extracted and summarised. Findings suggest that the popularity of modelling within this field is increasing over time with most studies published after 2015 and that population-based, deterministic, linear models were most frequently utilised, with the Lives Saved Tool being applied in over half of the reviewed studies (n=34, 57%). Much less frequently (n=6) models utilising system-thinking approaches, such as individual-based modelling or systems dynamics modelling, were developed and applied. Models were most applied to estimate the impact of interventions or services on maternal or neonatal mortality outcomes with morbidity-related outcomes and stillbirth reported on much less often. Going forward, given that healthcare delivery systems have long been identified as complex adaptive systems, modellers may consider the advantages of applying systems-thinking approaches to evaluate the impact of maternal and perinatal health policy. Such approaches allow for a more realistic and explicit representation of the systems- and individual-level factors which impact the effectiveness of interventions delivered within health systems.

## Introduction

The evaluation of the impact of healthcare programmes or policies on population health is a central component of public health, epidemiological and health economics science. Whilst the effects of biomedical interventions, such as drugs, on health outcomes are almost exclusively evaluated using randomised control trials (RCTs), experimental measurement of the direct effect of health policy interventions (e.g., changes to service coverage), may not be feasible due to ethical or practical constraints (1). In such instances, mathematical models, defined here as models which “set out a theoretical framework that represent the causal pathways and mechanisms linking exposures, interventions, and infection or disease”, can be utilised to estimate policy intervention effectiveness (1). Whilst the structure, assumptions and relative merits or limitation of such models varies considerably (1), the inherent flexibility of mathematical modelling, coupled with other intrinsic benefits such as lower relative cost when compared to experimentation, and the ability to synthesise multiple different data sources, has meant that they have come to be used extensively in the development of health policy at both the local governmental (2) and the global (3) levels.

Importantly, the application of mathematical models to inform health policy has the potential to generate significant health gains, particularly within the field of global maternal and newborn health. Evidence suggests that, despite significant progress since the turn of the century (4,5), maternal and perinatal mortality remains unacceptably high, particularly in regions such as southeast Asia and sub-Saharan Africa (SSA) (6–8), due in part to insufficient coverage and quality of key maternity services (9–14). As such, there are a substantial number of pertinent policy questions regarding how best to deliver high-quality maternity services to mothers and newborns to reduce mortality and maximise health which are of particular interest to policy makers within the region. This is especially true given the prominence of maternal and newborn health metrics within the health-related Sustainable Development Goals targets (15). For example, significant attention is now being paid to the potential health gains associated with maternity service delivery redesign (16,17), defined as the “intentional reorganisation of a health system to improve equity, quality, and outcomes” (17) which entails redesigning maternity services so that births occur exclusively within higher level or advanced facilities in line with practice in most high-income countries (16). Evaluation of such policies through experimentation, such as through a cluster RCT, is associated with substantial ethical constraints such as a potential lack of clinical equipoise. Additionally, certain outcomes of interest, such as maternal mortality, may be sufficiently rare that the required sample size is prohibitive to conducting a trial. In such circumstances, mathematical modelling may offer important insights into the potential impact of alternative configurations of intervention or service delivery.

Whilst there is evidence of the growing use of modelling approaches within public health more generally (18), to date there has been no systematic evaluation of the broad application of mathematical modelling to evaluate maternal and newborn services within the region of SSA. The review presented here has therefore been conducted to answer the primary research question of: *“How have mathematical models been used to evaluate the delivery of maternal and/or perinatal healthcare interventions or services and their impact on health-related outcomes in sub-Saharan Africa?”*. To best answer this question, the review has three primary objectives: (i) to characterise and map the relevant literature base; (ii) to explore the characteristics of the models and modelling studies found within this literature and (iii) to critically discuss the most common modelling approaches utilised. Systematic evaluation of the relevant literature allows for identification of common approaches taken, trends in study and model characteristics and potential gaps within the literature. Due to the broad scope of the research question and inclusion criteria we considered it appropriate to conduct a scoping review as opposed to a systematic review.

## Materials and methods

Levac et al’s. (19) framework was chosen to guide the conduct of this review as it provided the most comprehensive and clear guidance building upon the original six-stage framework for scoping review conduct developed by Arksey and O’Malley (20). This framework was utilised in tandem to the PRISMA-ScR checklist developed by the Joanna Briggs institute (21) to ensure the content of the review is in keeping with recommended best practice. The checklist for this review is provided in the supporting information (S1 File). We did not previously develop a review protocol.

### Inclusion criteria

Studies were eligible for inclusion if they described analyses using a mathematical model, as defined in the introduction, which reported the potential effect of the implementation or change in coverage of a maternal or perinatal health intervention on maternal or perinatal epidemiological or health related outcomes.

Maternal and perinatal healthcare interventions or services (referred to as interventions for the remainder of this review) are those interventions delivered as part of antenatal, intrapartum, or postpartum care to women at any stage along the pregnancy continuum (i.e., from conception to 42 days postnatal) or their newborns (i.e., from birth to day 28). For mothers, we only consider interventions intended to prevent morbidity or mortality associated with conditions or complications directly caused by the processes of pregnancy or birth.

Health outcomes include any measure of morbidity or mortality within the target population. This is not limited to the primary outcomes of the study and any studies which report outcomes partially derived from other health outcomes (e.g., Incremental Cost Effectiveness Ratio (ICER)) or that report health outcomes as secondary outcomes were included.

The geographical area of focus for this review was SSA with the full list of included countries being adapted from the UN Statistics Division geographic regions (22). Whilst the term SSA is a largely arbitrary categorisation of many extremely heterogenous and diverse countries, it was deemed a logical regional category as this grouping of African nations has been used extensively within the global health literature and therefore utilised to ensure the maximum number of relevant papers were returned.

Finally, papers in which mathematical models were described but not applied were not included, however those which used models previously applied in were. In addition, only English language papers were included in the review due to unavailable resources to undertake translation and/or non-English language searches.

### Search strategy

A multi-stage search strategy was employed to identify relevant studies for inclusion. First, a systematic search of three electronic databases was undertaken during November 2023. The databases included PubMed, Web of Science and Global Health (1973-present). Once relevant studies were identified, a hand-search of the bibliographies of selected studies was undertaken to identify any additional relevant sources of information. Table 1 presents the search terms used for searching the electronic databases. Due to the proliferation of studies identified in the first stage in which analyses were conducted using the Lives Saved Tool (LiST) (23), we undertook an additional search using the PubMed database using the search term “lives saved tool” to ensure all relevant studies were included in the final sample.

**Table 1.** Search terms used in database search of scoping review.

| Key term/concept | Search Terms |
| --- | --- |
| *Mathematical Model(s)* | ("mathematical" OR "disease" OR "microsimulation" OR "deterministic" OR "stochastic" OR "compartment*" OR "individual-based" OR "individual based" OR "agent-based" OR "agent based" OR "discrete-event" OR "time to event" OR "system dynamics" OR "monte carlo" OR "markov chain" OR "simulation") AND ("model*" OR "simulat*") |
| *Maternal and/or Perinatal Health* | "matern*" OR "pregnan*" OR "obstetric*" OR "antenatal care" OR "prenatal care" OR "postnatal care" OR "postpartum care" OR "birth attendan*" OR "intrapartum care" OR "labour and delivery" OR "facility delivery" OR "child birth" OR "childbirth" OR "neonat*" OR "perinatal" OR "newborn" OR "new born" OR "BEmONC" OR "EmONC" OR "CEmONC" OR "emergency obstetric and newborn care" OR "emergency obstetric and new born care" |
| *Sub-Saharan Africa* | Angola* OR Benin OR Botswana* OR "British Indian Ocean Territory" OR "Burkina Faso" OR Burundi* OR "Cabo Verde" OR Cameroon* OR "Central African Republic" OR Chad OR Congo OR Comoros OR "Cote d'Ivoire" OR "Democratic Republic of the Congo" OR Djibouti* OR Eswatini* OR Eritrea* OR Ethiopia* OR "equatorial guinea" OR "French Southern Territories" OR Gabon OR Gambia* OR Ghana* OR guinea OR "Guinea-Bissau" OR Kenya* OR Lesotho OR Liberia* OR Madagascar* OR Malawi* OR Mali* OR Mauritania* OR Mauriti* OR Mayotte OR Mozambi* OR Namibia* OR Niger* OR Nigeria* OR "Saint Helena" OR Reunion OR Rwanda* OR "Sao tome and Principe" OR Senegal* OR Seychell* OR "Sierra Leon*" OR Somalia* OR "South Africa*" OR "South Sudan" OR Tanzania* OR Togo* OR Uganda* OR Zambia* OR Zimbabwe* OR "sub-Saharan Africa" |

Citations returned from each database search were imported into Mendeley Desktop citation manager (v1.19.8) and pooled. Using in-built functionality automatic deduplication was undertaken followed by a manual review of returned studies to ensure any duplicates missed by the automatic search were removed.

### Study selection

A three-step process was employed to identify relevant studies from retrieved titles; 1.) the titles of the total number of retrieved citations were screened for apparent relevance using the inclusion criteria, 2.) the potentially relevant studies were separated then screened by the contents of their abstract against the eligibility criteria, 3.) those which still appeared relevant were screened through review of the full text and included in the review if appropriate. This process was mirrored with any studies identified via the hand-search of the reference lists of the final studies selected from the database and those identified through the targeted search. Despite recommendations within current methodological frameworks that this process, and the process of extraction and analysis, should be conducted by a multidisciplinary team to improve the validity of the review’s findings (19,21,24,25), this review was initially conducted as part of the first authors Ph.D. and therefore was conducted with no additional authors.

### Data extraction and analysis

Following the selection of relevant studies, data was abstracted from the final studies via a data abstraction form. The development and use of a unique abstraction form is central to scoping review methodology and allows researchers to identify and extract all pertinent information from a given study which relates directly to the reviews primary research question (19,26). As such a preliminary data extraction form was first developed and trialled on a subset of five studies, in-keeping with recommended scoping review practice (19,26), however due to the significant heterogeneity in study methods and analyses a second round of trialling was conducted on a further five studies after the initial refinement of the data extraction form. Within the final form 35 unique data items are identified. They can broadly be categorised as general characteristics of the study (8 items), characteristics of modelled interventions (9 items), characteristics of the mathematical model (7 items) and characteristics of any conducted analyses (11 items). The final completed data extraction form can be found in the supporting information (S2 File).

Data items relating to model characteristics were adapted from Garnett et al. (1), where the authors provide defining criteria relevant across different mathematical modelling methodologies. These criteria, which have been refined for data extraction, include how feedback between interventions and outcomes are represented in the modelled system, the role chance plays in model behaviour, how individuals are represented within the model and how variable change is governed with regards to time (1).

Following data extraction, data items were synthesised quantitatively where appropriate (i.e., in the presentation of percentages and frequencies) and summarised under relevant subheadings presented in the following section.

## Results

### Search results

A total of 10,980 potentially relevant studies were identified, which was reduced to 8,660 after the removal of duplicates. Following title screening, the abstracts of 150 papers were screened (including all papers from hand-searching) using the inclusion criteria followed by full-text screening of 90 most relevant. Of these studies a further 30 were excluded leaving a final total of 60 papers for review (27–86). Figure 1 provides a diagrammatic representation of the search and screening processes within this review.

**Figure 1.**
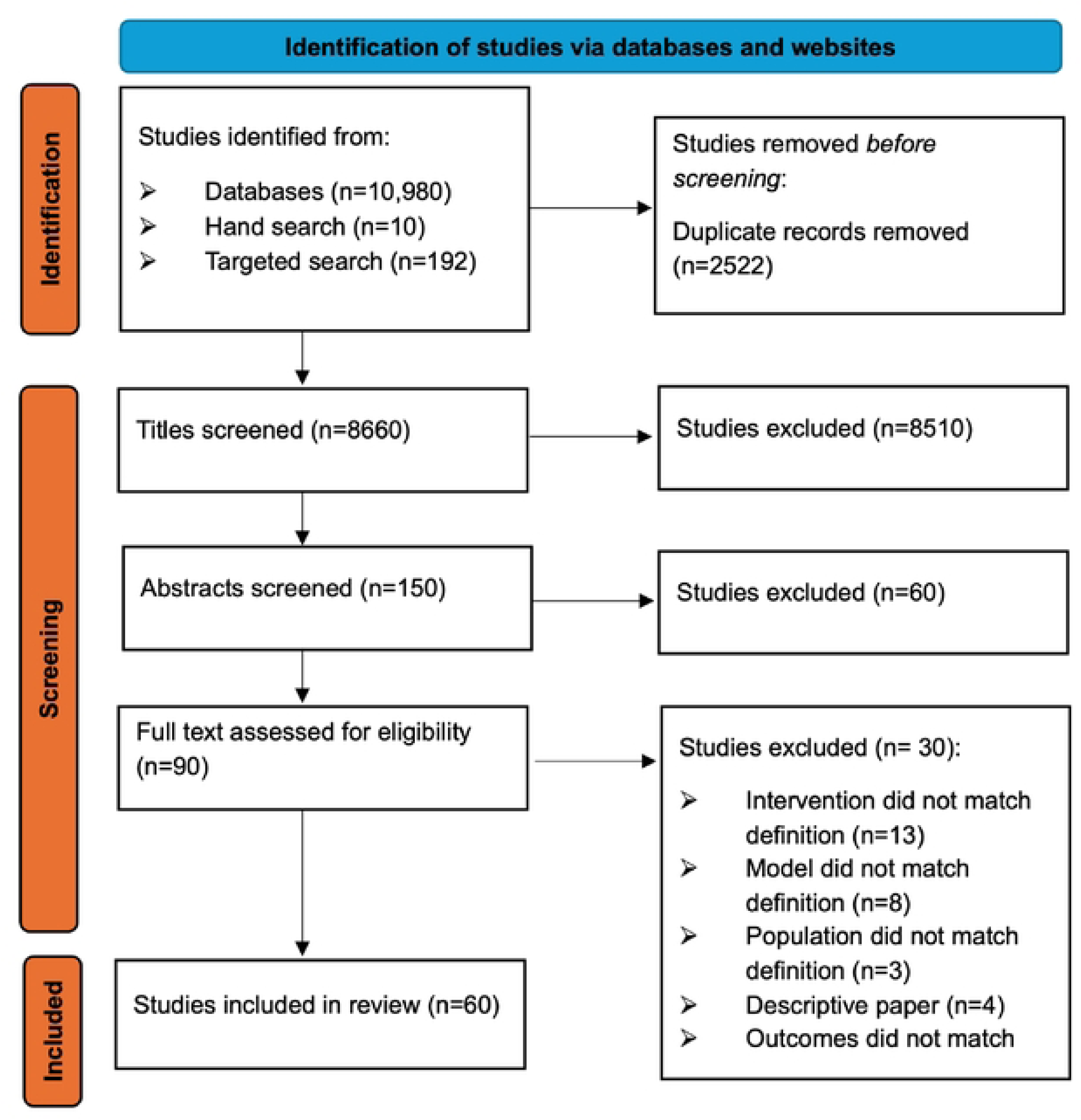
PRISMA diagram reporting search results.

### General study characteristics

Table 2 details the general characteristics of the included studies. The publication date of the included studies skewed towards the latter half of the 2010s, with 70% of studies published after 2015 (n=42) and the greatest number of studies being published in 2019 (n=9 (15%)). In over half of studies (n=33, 55%) the modelled ‘population’ was representative of a single country, whilst the remaining studies either modelled interventions applied to populations at a regional level, such as sub-Saharan Africa (n=8), or to two or more countries or regions (n=19).

**Table 2.** General characteristics of the reviewed studies.

| **General study characteristics** | **Studies**<br>**(N=60)** | **Percentage**<br>**(%)** |
| --- | --- | --- |
| *Year of publication:* |  |  |
| Pre 2010 | **4** | **7%** |
| 2010-2014 | **14** | **23%** |
| 2015-2019 | **34** | **57%** |
| 2020-2023 | **8** | **13%** |
| *Single or multiple countries modelled:* |  |  |
| Single | **33** | **55%** |
| Multiple | **27** | **45%** |
| *Most modelled countries (top 5) <sup>a</sup>* |  |  |
| Ethiopia | **21** | **35%** |
| Tanzania | **19** | **32%** |
| Uganda | **18** | **30%** |
| Kenya | **18** | **30%** |
| Malawi | **16** | **27%** |
| *In addition to health outcomes the study aimed to evaluate cost/cost-effectiveness:* |  |  |
| Yes | **22** | **37%** |
| No | **38** | **63%** |
| *First author affiliation:* |  |  |
| Within study country | **18** | **30%** |
| Outside study country | **42** | **70%** |
| *Any author based at institution within the/a study country:* |  |  |
| Yes | **31** | **52%** |
| No | **29** | **48%** |

<sup>a</sup>Sourced from studies which explicitly listed modelled countries. Additionally, multiple countries are modelled in many papers therefore values in this row when summed exceed the total number of papers

Location of the affiliated institution of the first study author was extracted, alongside whether any co-authors were based in institutions within study countries. Seventy percent (n=40) of studies’ first authors institutions were not located in any of the study countries with nearly half of total studies led by a first author based at an institution in the United States (n=29, 48%) and only seventeen studies (28%) were led by first authors based at institutions within SSA. As evident in Table 2, approximately half of studies (n=29) had no authors based at an affiliated institution within any of the study countries.

Finally, the primary aim of included studies was evaluated. Whilst all studies reported at least one outcome associated with either maternal or perinatal health in line with inclusion criteria, over a third (n=22, (37%)) aimed to estimate costs associated with modelled interventions or calculate the interventions’ cost-effectiveness.

#### Intervention characteristics

Table 3 summarises extracted data relating to the interventions evaluated within the reviewed studies. Interventions were extracted, as written, from either within the main text or supplementary material if indicated by authors. They were then alphabetised to assess frequency and, where wording differed slightly between interventions which could reasonably be considered the same, this was corrected.

**Table 3.** Characteristics of the interventions included within the reviewed studies.

| Intervention characteristics | Studies<br>(N=60)<sup>a</sup> | Percentage (%) |
| --- | --- | --- |
| Study evaluated a single or multiple interventions: |  |  |
| *Single* | 8 | 13% |
| *Multiple* | 52 | 87% |
| Study modelled an intervention not currently delivered as part of routine service delivery: |  |  |
| Yes | 7 | 12% |
| No | 53 | 88% |
| Study includes interventions delivered to: |  |  |
| *Women* | **57** | 95% |
| *Neonates* | **41** | 69% |
| If a study modelled any interventions delivered to mothers, they were delivered: |  |  |
| *Antenatally* | **47** | 78% |
| *During labour/birth* | **43** | 72% |
| *Postnatally* | **41** | 68% |
| Study modelled interventions targeting: |  |  |
| *Maternal health outcomes* | **42** | 70% |
| *Neonatal health outcomes* | **46** | 77% |
| *Stillbirths* | **17** | 28% |
| Most evaluated interventions (Top 5)<sup>b</sup>: |  |  |
| 1. *Immediate assessment and stimulation / Neonatal resuscitation* | **34** | 57% |
| 2. *Skilled birth attendance* | **32** | 53% |
| 3. *Vitamin A supplementation* | **24** | 40% |
| 4. *Clean birth or postnatal practices* | **24** | 40% |
| 5. *Magnesium Sulphate* | **20** | 33% |

<sup>a</sup>The values in the some of the rows exceed the total number of papers when summed, as most studies modelled multiple interventions.
<sup>b</sup>Excluding those interventions which are not routinely delivered as part of antenatal, intrapartum, or postpartum care

There was extensive heterogeneity between studies in relation to the interventions which were evaluated by mathematical models and the ways in which interventions were defined. Across the 60 included studies a total of 1055 interventions (inclusive of duplicates) were described, with most studies (n=52 (87%)) evaluating the impact of multiple different interventions and fewer studies evaluating single interventions. This value includes all modelled interventions including those which are delivered outside routine antenatal, intrapartum, or postpartum care or represented nonclinical interventions such as improved access to clean water.

Commonly, studies evaluated scale-up of interventions which could reasonably be defined as part of standard clinical practice in most settings (e.g., basic newborn resuscitation). Much less frequently, (n=7, 12%) studies evaluated novel interventions which are not routinely available to mothers or newborns in most settings. This includes studies in which the aim was to predict the effect of an intervention on health outcomes that acts through improving coverage of other key maternity services. For example, Wilcox et al. (28) predicted the impact of a Mobile Technology for Community Health initiative in Ghana which has been shown to improve skilled birth attendance and facility delivery and modelled the effect of this intervention using LiST. Other examples include Carvalho et al. (55) in which the authors estimated the cost-effectiveness and health impacts of an inhaled oxytocin product and Ali et al. (58) who evaluated the Augmented Infant Resuscitator, a novel device which provides real-time feedback on ventilation quality during the resuscitation of newborns.

Almost all studies evaluated at least one intervention which was delivered to women (N=57 (95%)), even including those in which maternal outcomes were not evaluated or reported (e.g., evaluating the effect of maternal nutritional supplementation in ANC on newborn outcomes). As shown in Table 3 these interventions were most frequently delivered during the antenatal and intrapartum periods of pregnancy whilst postpartum interventions delivered to mothers were less frequent (n=41, 68%). The evaluation of interventions delivered to neonates occurred slightly more frequently and was presented in over two-thirds of studies (n=41 (68%)). Of the studies in which only neonatal interventions were evaluated (n=2), Tsiachristas et al. (29) evaluated a task shifting intervention for routine nursing tasks delivered to neonates in a Kenyan neonatal intensive care unit (NICU) whilst Ali et al. (58) has been described above.

### Model characteristics

Table 4 summarises the extracted data relating to the models applied within the reviewed studies. Approximately a quarter (n=16 (27%)) of studies utilised a model which had been developed specifically for the purpose of achieving one or more of the study’s aims whilst the remaining employed models which had been developed prior to study conduct. Over half of studies (n=34, 57%) reported analyses conducted using LiST, and eight studies (13%) using the Maternal and Neonatal Directed Assessment of Technology model (MANDATE).

**Table 4.** Characteristics of the models within the reviewed studies.

| Model characteristics | Studies (N=59) | Percentage (%) |
| --- | --- | --- |
| Was the model developed to conduct this study: |  |  |
| Yes | 16 | 27% |
| No | 44 | 73% |
| Model employed by study authors: |  |  |
| *LiST* | 34 | 57% |
| *Maternal and Neonatal Directed Assessment of Technology model (MANDATE)* | 8 | 13% |
| *Decision Tree/Analytic model* | 7 | 12% |
| *Individual or Agent-based model* | 5 | 8% |
| *Systems-dynamics model* | 1 | 2% |
| *Other/Undefined* | **5** | **8%** |
| Model structure, use or data and reporting: |  |  |
| *System feedback – Linear* | **58** | **97%** |
| *System feedback – Non-linear* | **2** | **3%** |
| *Deterministic* | **55** | **92%** |
| *Stochastic* | **5** | **8%** |
| *Population/cohort based.* | **55** | **92%** |
| *Individual-based* | **5** | **8%** |
| *Time represented as calendar time.* | **53** | **88%** |
| *Time represented with age* | **7** | **12%** |
| *Diagrammatic representation in manuscript* | **13** | **22%** |
| *No diagrammatic representation in manuscript* | **47** | **78%** |

Of the remaining studies in the sample, most utilised a decision tree/analytic model (n=7). These models, commonly used in health economic analysis, represent potential treatment pathways as a series of ‘branches’ demarcated by decision nodes, representing some decision of interest (e.g., the decision to deliver a screening intervention) and chance nodes from which mutually exclusive probabilities of a set of outcomes following a decision are represented (87,88). Other types of models were utilised much less frequently with only 5 (8%) studies employing an individual-based framework (27,32,34,68,86).

Model characteristics were extracted from all included studies guided in part by Garnett et al. (1). Considering whether the link between a given intervention and the health outcome in the model is represented as a linear or non-linear function, 95% of models can be described as linear (n=57). A simple linear model would assume that if a given treatment halves the risk of disease acquisition, and that treatment is delivered to everyone at risk of the condition, then the incidence of that condition would reduce by half in the population. Linear representations are common in cohort models where it is assumed that the outcome of different individuals represented in the model is independent (i.e., treatment delivered or not delivered within the model to one individual does not affect the treatment or outcomes of others). The fact that most models reviewed here are linear is unsurprising due to the number of studies using either LiST or the MANDATE model which both are self-described as linear. Alternatively, models which represent this link as non-linear functions were much less common (n=2) (31,34).

Next, considering the incorporation of chance into the modelling framework, few models (n=5, 8%) were found to be stochastic (i.e., events occur to an individual or population group within the model according to random draws against a probability parameter), while most utilised a deterministic approach to derive model results, meaning that multiple model runs with the same parameter set will always return the same result (n=55). All stochastic models within the review employed an individual-based approach, where each member of the population is modelled individually with distinct characteristics which can be tracked, whilst the remaining models included representation of population level processes and trends using input parameters representing averages and leading to population-level results.

Description of model structure and components varied according to model type. Only 13 (22%) papers presented any diagrammatic representation of the model’s underlying structure within the manuscript or supplementary material. However, of the papers without a model diagram over 80% of these papers utilised either LiST or MANDATE model. Diagrammatic representation of model structure is available for both these models on their associated websites (89,90). This meant that papers in which novel models were reported were significantly more likely to include a diagram of the structure.

### Study analyses and outcomes

Table 5 is a broad overview of the analyses and relevant outcomes extracted from the reviewed studies. All study outcomes were extracted, as presented, from each study and then sorted according to categories presented in Table 5. Of note, cost related outcomes are not included in the table but are available in the supporting information (S2 File). Maternal and neonatal mortality outcomes were by far the most frequently reported within the sample with very few studies reporting outcomes which could reasonably be classified as relating to maternal (n=4, 7%) or neonatal (n=6, 10%) morbidity. Interestingly, of the studies presenting maternal morbidity outcomes (38,45,55,61) all reported some measure of postpartum haemorrhage incidence or prevalence whilst newborn morbidity outcomes ranged from the prevalence of stunting or wasting (56) to the number of cases of intrapartum related neonatal events and injury (43). Additionally, any measure of the number or rate of stillbirths within a modelled population was reported in only around a fourth of studies within the sample (n=14, 23%).

**Table 5.** Characteristics of the analyses in the reviewed studies.

| Characteristics of analyses | Studies<br>(N=59) | Percentage<br>(%) |
| --- | --- | --- |
| (Where relevant (n=53)) Calendar-time horizon for analysis was: |  |  |
| *One year or less* | **20** | 34% |
| *2-5 years* | **11** | 18% |
| *6-10 years* | **5** | 8% |
| *11+ years* | **17** | 28% |
| (Where relevant (n=7)) Age-time horizon for analysis was: |  |  |
| *A delivery* | **3** | 5% |
| *A pregnancy and delivery* | **1** | 2% |
| *2 years* | **1** | 2% |
| *Lifetime* | **2** | 3% |
| Sensitivity analysis was conducted and reported on: |  |  |
| Yes | **20** | 33% |
| No | **40** | 67% |
| Study reported outcome relating to: |  |  |
| *Maternal mortality* | **34** | 57% |
| *Maternal morbidity* | **4** | 7% |
| *Maternal DALYs* | **6** | 10% |
| *Neonatal mortality* | **39** | 65% |
| *Neonatal morbidity* | **6** | 10% |
| *Neonatal DALYs* | **7** | 12% |
| *Stillbirths* | **14** | 23% |
| For multi-country studies (n=27) results were reported by country or as an aggregate over a region/group? |  |  |
| *Country-specific* | **9** | 33% |
| *Aggregate* | **18** | 67% |

Additionally, we considered how outcomes were presented for studies in which more than one country population was theoretically modelled. Surprisingly, of the multi-country studies (n=27), only eight reported results from their analysis disaggregated by each included country, with the rest presenting the results for a group of countries or even an entire region, such as SSA. Whilst for some studies this was an analytical choice, for those using the MANDATE model authors were limited by the design of the model which currently only represents the region of SSA or the country of India within the model’s base assumptions (e.g., condition incidence, intervention availability).

## Discussion

This scoping review has provided a comprehensive and descriptive overview of mathematical modelling within maternal and perinatal health research within SSA. Through application of Levac et al’s. (19) framework and guided by the recent PRISMA-ScR checklist (21), a replicable evaluation of the published academic literature has been undertaken in which the characteristics of relevant mathematical models and their application in the evaluation of maternal and/or perinatal interventions has been summarised. Given that the number of studies in the field utilising these approaches appears to have increased over time, as evidenced in this review, this is a timely and pertinent overview of commonly utilised modelling approaches. The proliferation of studies from 2015 onwards could be attributed to several factors including the utility of such methods in evaluating pathways towards achieving maternal and perinatal health related goals within the Millennium Development Goals (MDGs) and Sustainable Development Goals (SDGs), as demonstrated in several reviewed studies (79–85).

Our review found that the most often utilised model within this field was LiST, which was used in nearly sixty percent of the sample. LiST has long been recognised as a popular tool which has been utilised extensively within the field of global maternal and child health by a varied group of interdisciplinary stakeholders (91). LiST is a linear deterministic model as it “describes fixed relationships between inputs and outputs, and the tool will produce the same outputs each time the model is run with identical inputs” (92). The primary input is the coverage of an intervention being evaluated and the primary outputs are either change in the cause-specific mortality or one of the included “risk factors” for mortality such as stunting (92). LiST is extremely comprehensive in both the breadth of causes of maternal and child ill-health which are represented and the range of in-built interventions which modellers can manipulate within the tool and has been utilised in the evaluation of health programmes and to inform strategic planning at the national and local level with some organisations regularly using the tool and embedding LiST into organisational workflows by developing ‘in-house’ capacity to work with the model (91). Importantly, as demonstrated in Table 4, many of the other models in this sample share key characteristics with LIST in that they are linear, deterministic, and changes in the modelled system are expressed at the population level.

However, healthcare delivery systems have long been identified as complex adaptive systems which exhibit explicitly non-linear behaviour by definition (93–95). Variation in the coverage of established interventions, or integration of new interventions, may have unexpected down-stream effects on the outcome of interest despite evidence that the intervention is effective (96,97) which cannot be predicted using LiST or similar linear models. Within maternal and perinatal health this has been demonstrated in several Low- and middle-income countries (LMICs) in which, driven by political commitment to achieving the maternal health indicators within MDG 5 and SDG 3, the proportion of mothers who give birth within a health facility has increased significantly (96,97). However, this has not been accompanied by an observed population-level reduction in maternal mortality, despite strong evidence for the efficacy of interventions delivered to mothers during and following labour (96,97). The relationship between facility delivery coverage and mortality is complex, non-linear, and likely influenced by several individual and health system-level factors relating to quality of care such as the effect of increased volume of patient attendance and resource use (96,97). Failure to understand the system environment and behaviours is often a driving factor in the failed implementation of new interventions, which may have been effective had a systems-thinking lens, one which considers the potential effect of system level phenomena on intervention or service implementation, been employed (98–100). This issue is further compounded when the complexity of the interventions is increased, as many health systems strengthening interventions are largely programmatic service changes which mean current evaluation methodologies may be insufficient to produce valid or reliable outcomes (100).

Evaluation of relevant health processes at the population level, as taken by most models in this review, is advantageous in that it is computationally cheap and, in many circumstances, may be a sufficient level of detail to ascertain a reliable result (101). However, in recent years, the use of individual-based model (IBMs) has become more popular within epidemiology and public health (18,102,103). Broadly, IBMs are a subset of computational simulation models in which the model is designed to replicate the characteristics, behaviours, and actions of a set of ‘agents’, within a given environment, to reproduce complex population-level or systems phenomena (104–107). Within epidemiological modelling, these ‘agents’ are often representative of individual people who may interact with one another to simulate a hypothetical population of interest, characterised by individual variables, and governed by sets of predetermined rules (105).

One of the primary strengths of this approach in the modelling of health processes is the ability to explicitly model individual-level variation and interactions between said individuals within the population (104). Within epidemiological modelling, this approach has been employed in the modelling of infectious diseases, in which interactions between individuals allows for representation of complex transmission dynamics within the population (103). This process, often referred to as ‘interference’, occurs because individuals within the model are not represented as independent, and therefore the assignment of an exposure, intervention or treatment can be influenced by outcomes of other individuals in the model (105). Interference cannot be represented in population-level models where the assumption of non-independence is not met which means such models are less suitable to explore the complexities of healthcare delivery. This is demonstrated in the study by Nadkarni et al. (34), who explicitly represent the relationship between the acuity of a group of patients in a maternity ward, the associated demand on consumables and, in-turn, the effect on outcomes.

Our review found that only 27% of research teams developed novel mathematical models to answer the research question of interest. Whilst the appeal of using a previously developed model is clear, the importance of considering country-context within the evaluation of health policy has long been identified within the literature, as national and regional context likely has significant impact on how interventions are implemented and accessed by populations (108). Many modellers have embraced methods to embed contextual structures into model design through participatory approaches to model development (109), stakeholder workshops developing causal loop diagrams (110), or simply through explicitly representing the system of interest within a given context by supported model validation with key stakeholders (111). Whilst such approaches were underutilised in the reviewed studies, context-informed modelling was well demonstrated by Semwanga et al. (31) who developed a novel systems-dynamics model to explore neonatal health interventions in Uganda through a process of stakeholder engagement and causal mapping with service users and policy makers.

Next, considering the commonly reported outcomes of modelling analyses, this review found that studies were considerably more likely to report outcomes relating to either maternal and neonatal mortality and much less likely to predict intervention effects on morbidity or stillbirths. Limited reporting on morbidity outcomes is not surprising due to the widespread use of LiST which does not calculate the effect of interventions on most morbidity outcomes. The absence of morbidity from the outcomes of modelling studies evaluating health intervention delivery is concerning considering that many of the routine interventions delivered to mothers and newborns are intended not only to prevent mortality but to reduce the incidence of common morbidity associated with pregnancy and the newborn period to improve overall quality of life (112–114). The absence of such outcomes may mean that the true health impact of improving intervention coverage is significantly underestimated at the population level.

Finally, we found that very few of the included studies evaluated the potential impact of interventions on stillbirths. There are similarities between stillbirth research and morbidity research, with many arguing that stillbirth has historically been absent from the global maternal and perinatal health agenda for some time (115–117). Qureshi et al. (117) suggest that stillbirths should be attributed the same ‘value’ as neonatal deaths in economic evaluation of maternal health interventions as they are commonly excluded in measurements such as life years gained or DALYs averted. This is problematic when considering very early preterm neonatal deaths are included in neonatal death statistics whilst a stillbirth at 41 weeks gestational age would not be, despite being more developmentally ‘mature’ (117). Whilst recently a renewed focus on stillbirth prevention has led to considerable progress (118) a large number of stillbirths occur per year within SSA (6) and are associated with considerable morbidity in mothers (119). Importantly several interventions to prevent stillbirth are well described in the literature through a Cochrane review of relevant trials (120) and through observational studies (121). As such mathematical modelling approaches are well positioned to evaluate the context-specific effects of these interventions in high burden populations.

There are several important strengths and limitations of this review to acknowledge. The primary strength of this review is the use of a systematic search strategy to identify relevant studies and due to the nature and broad scope of the research question and associated objectives, the scoping review methodology was the most appropriate choice to map the relevant literature. A traditional systematic review would not have been appropriate, as the aim of this review was not to synthesise the reported results of the modelling studies, something that would not have been possible due to the diverse set of methodologies employed within this research. To ensure that conduct of the review adhered to contemporary best-practice frameworks the PRISMA checklist (21) was used to guide the design and conduct of the review (S1 File).

Considering this review’s limitations, firstly the search strategy did not include modelling analyses reported within the grey literature. Implicit exclusion of potentially relevant modelling analyses conducted in reports or government documentation could introduce bias into the review and potentially alter the review’s conclusions (122). Interestingly, the LiST team highlights in their most recent update supplement that many applications of the model lie outside of academic publishing, with efforts being made by the team to enhance the visibility of LiST applications by policy makers across LMICs (123). Whilst frameworks for the systematic search of grey literature do exist, they do not alleviate common issues with comprehensive grey literature searching such as inbuilt search-engine filters which can introduce bias into source selection (124).

Secondly, a significant number of modelling studies which evaluated solely the impact of health interventions on indirect causes of maternal morbidity/mortality were excluded. This was particularly true for modelling studies evaluating screening and treatment of HIV during pregnancy on HIV outcomes. Undoubtedly, widening the studies inclusion criteria so that such studies were reviewed would likely have changed the results of this review. This is particularly true given the proliferation of HIV models used to conduct research in SSA (125,126). Finally, this review did not explicitly consider the quality of included studies by conducting a critical appraisal as part of the review process. Whilst evaluation of the quality of modelling studies conducted in this area is undoubtedly important, this process is outside of the remit of a scoping review (19,127) and is beyond the objectives of this review.

In conclusion, in this review we have reported on the methodology, conduct and results of a scoping review to explore the breadth of mathematical modelling research within evaluation of maternal and perinatal healthcare interventions in SSA. Results from this review indicate that whilst there is variation in the reviewed studies, the literature base is dominated by two models which, whilst distinct in their design, share commonalities in how they represent the relationship between intervention and outcome, populations of interest and the role randomness plays in the model system. Individual-based or systems-thinking approaches to mathematical modelling were much less commonly applied within the field, despite potential advantages such as a more realistic and explicit representation of the systems- and individual-level factors that impact the effectiveness of interventions delivered within complex-adaptive health systems.

## Data Availability

All relevant data are within the manuscript and its Supporting Information files.

## Supporting information

S1 File. PRISMA-ScR Checklist

S2 File. Data extraction form and extracted data

## References

1. Garnett G, Cousens S, Hallett T, Steketee R, Walker N. Mathematical models in the evaluation of health programmes. The Lancet. 2011;378(9790):515–25. Available from: https://pubmed.ncbi.nlm.nih.gov/21481448/

2. Panovska-Griffiths J, Kerr CC, Waites W, Stuart RM. Mathematical modeling as a tool for policy decision making: Applications to the COVID-19 pandemic. Handbook of Statistics. 2021;44:291–326. Available from: https://www.ncbi.nlm.nih.gov/pmc/articles/PMC7857083/

3. Lo NC, Andrejko K, Shukla P, Baker T, Sawin VI, Norris SL, et al. Contribution and quality of mathematical modeling evidence in World Health Organization guidelines: A systematic review. Epidemics. 2022;39:1–8. Available from: https://pubmed.ncbi.nlm.nih.gov/35569248/

4. World Health Organization, UNICEF, UNFPA, World Bank Group, UNDESA/Population Division. Trends in maternal mortality 2000 to 2020. 2023. Available from: https://www.who.int/publications/i/item/9789240068759

5. Kassebaum NJ, Barber RM, Bhutta ZA, Dandona L, Gething PW, Hay SI, et al. Global, regional, and national levels of maternal mortality, 1990–2015: a systematic analysis for the Global Burden of Disease Study 2015. The Lancet. 2016;388(10053):1775–812. Available from: https://linkinghub.elsevier.com/retrieve/pii/S0140673616314702

6. Hug L, You D, Blencowe H, Mishra A, Wang Z, Fix MJ, et al. Global, regional, and national estimates and trends in stillbirths from 2000 to 2019: a systematic assessment. The Lancet. 2021;398(10302):772–85. Available from: http://www.thelancet.com/article/S0140673621011120/fulltext

7. Kassebaum NJ. Global, regional, and national progress towards Sustainable Development Goal 3.2 for neonatal and child health: all-cause and cause-specific mortality findings from the Global Burden of Disease Study 2019. The Lancet. 2021;398(10303):870–905. Available from: https://pubmed.ncbi.nlm.nih.gov/34416195/

8. Alkema L, Chou D, Hogan D, Zhang S, Moller AB, Gemmill A, et al. Global, regional, and national levels and trends in maternal mortality between 1990 and 2015, with scenario-based projections to 2030: A systematic analysis by the UN Maternal Mortality Estimation Inter-Agency Group. The Lancet. 2016 Jan 30;387(10017):462–74. Available from: http://www.thelancet.com/article/S0140673615008387/fulltext

9. Andegiorgish AK, Elhoumed M, Qi Q, Zhu Z, Zeng L. Determinants of antenatal care use in nine sub-Saharan African countries: a statistical analysis of cross-sectional data from Demographic and Health Surveys. BMJ Open. 2022;12(2):e051675. Available from: https://bmjopen.bmj.com/content/12/2/e051675

10. Adde KS, Dickson KS, Amu H. Prevalence and determinants of the place of delivery among reproductive age women in sub–Saharan Africa. PLoS One. 2020;15(12). Available from: https://journals.plos.org/plosone/article?id=10.1371/journal.pone.0244875

11. Tessema ZT, Yazachew L, Tesema GA, Teshale AB. Determinants of postnatal care utilization in sub-Saharan Africa: a meta and multilevel analysis of data from 36 sub-Saharan countries. Ital J Pediatr. 2020;46(1):1–11. Available from: https://ijponline.biomedcentral.com/articles/10.1186/s13052-020-00944-y

12. Wigley AS, Tejedor-Garavito N, Alegana V, Carioli A, Ruktanonchai CW, Pezzulo C, et al. Measuring the availability and geographical accessibility of maternal health services across sub-Saharan Africa. BMC Med. 2020;18(1):1–10. Available from: https://bmcmedicine.biomedcentral.com/articles/10.1186/s12916-020-01707-6

13. Rosser JI, Aluri KZ, Kempinsky A, Richardson S, Bendavid E. The Effect of Healthcare Worker Density on Maternal Health Service Utilization in Sub-Saharan Africa. Am J Trop Med Hyg. 2022;106(3):939–44. Available from: https://pubmed.ncbi.nlm.nih.gov/35026729/

14. Kruk ME, Leslie HH, Verguet S, Mbaruku GM, Adanu RMK, Langer A. Quality of basic maternal care functions in health facilities of five African countries: an analysis of national health system surveys. Lancet Glob Health. 2016;4(11):e845–55. Available from: http://www.thelancet.com/article/S2214109X16301802/fulltext

15. The United Nations. Transforming our world: The 2030 Agenda for Sustainable Development. 2015. Available from: https://www.unfpa.org/resources/transforming-our-world-2030-agenda-sustainable-development

16. Roder-DeWan S, Madhavan S, Subramanian S, Nimako K, Lashari T, Bathula AN, et al. Service delivery redesign is a process, not a model of care. BMJ. 2023;380:e071651. Available from: https://www.bmj.com/content/380/bmj-2022-071651

17. Roder-Dewan S, Nimako K, Twum-Danso NAY, Amatya A, Langer A, Kruk M. Health system redesign for maternal and newborn survival: rethinking care models to close the global equity gap. BMJ Glob Health. 2020;5(10):e002539. Available from: https://gh.bmj.com/content/5/10/e002539

18. Cassidy R, Singh NS, Schiratti PR, Semwanga A, Binyaruka P, Sachingongu N, et al. Mathematical modelling for health systems research: A systematic review of system dynamics and agent-based models. BMC Health Serv. Res.. 2019;19. Available from: https://bmchealthservres.biomedcentral.com/articles/10.1186/s12913-019-4627-7

19. Levac D, Colquhoun H, O’Brien KK. Scoping studies: Advancing the methodology. Implement Sci. 2010;5(1):1–9. Available from: https://implementationscience.biomedcentral.com/articles/10.1186/1748-5908-5-69

20. Arksey H, O’Malley L. Scoping studies: towards a methodological framework. Int J Soc Res Methodol. 2005;8(1):19–32. Available from: https://pubmed.ncbi.nlm.nih.gov/25034198/

21. Tricco AC, Lillie E, Zarin W, O’Brien KK, Colquhoun H, Levac D, et al. PRISMA extension for scoping reviews (PRISMA-ScR): Checklist and explanation. Ann Intern Med. 2018;169(7):467–73. Available from: https://pubmed.ncbi.nlm.nih.gov/30178033/

22. UN Statistics Division. Standard country or area codes for statistical use (M49). 2023. Available from: https://unstats.un.org/unsd/methodology/m49/

23. Johns Hopkins Bloomberg School of Public Health, Bill & Melinda Gates Foundation. The Lives Saved Tool - LiST in peer-reviewed journals. 2022. Available from: https://www.livessavedtool.org/list-in-peerreviewed-journals

24. Peters M, Godfrey C, McInerney P, Munn Z, Trico A, Khalil H. Chapter 11: Scoping Reviews. In: Aromataris E, Munn Z, editors. JBI Manual for Evidence Synthesis, JBI 2020. Available from: https://jbi-global-wiki.refined.site/space/MANUAL/4687342/Chapter+11%3A+Scoping+reviews

25. Peters M, Godfrey C, Khalil H, McInerney P, Parker D, Soares CB. Guidance for conducting systematic scoping reviews. Int J Evid Based Healthc. 2015;13(3):141–6. Available from: https://pubmed.ncbi.nlm.nih.gov/26134548/

26. Pollock D, Peters MDJ, Khalil H, McInerney P, Alexander L, Tricco AC, et al. Recommendations for the extraction, analysis, and presentation of results in scoping reviews. JBI Evid Synth. 2022;21(3):520–532. Available from: https://pubmed.ncbi.nlm.nih.gov/36081365/

27. Young N, Bowman A, Swedin K, Collins J, Blair-Stahn ND, Lindstedt PA, et al. Cost-effectiveness of antenatal multiple micronutrients and balanced energy protein supplementation compared to iron and folic acid supplementation in India, Pakistan, Mali, and Tanzania: A dynamic microsimulation study. PLoS Med. 2022;19(2):e1003902. Available from: https://dx.plos.org/10.1371/journal.pmed.1003902

28. Willcox M, Moorthy A, Mohan D, Romano K, Hutchful D, Mehl G, et al. Mobile Technology for Community Health in Ghana: Is Maternal Messaging and Provider Use of Technology Cost-Effective in Improving Maternal and Child Health Outcomes at Scale? J Med Internet Res. 2019;21(2). Available from: https://pubmed.ncbi.nlm.nih.gov/30758296/

29. Tsiachristas A, Gathara D, Aluvaala J, Chege T, Barasa E, English M. Effective coverage and budget implications of skill-mix change to improve neonatal nursing care: an explorative simulation study in Kenya. BMJ Glob Health. 2019;4(6):e001817. Available from: https://gh.bmj.com/lookup/doi/10.1136/bmjgh-2019-001817

30. Stenberg K, Watts R, Bertram MY, Engesveen K, Maliqi B, Say L, et al. Cost-Effectiveness of Interventions to Improve Maternal, Newborn and Child Health Outcomes: A WHO-CHOICE Analysis for Eastern Sub-Saharan Africa and South-East Asia. Int J Health Policy Manag. 2021;10(11):706–23. Available from: https://pubmed.ncbi.nlm.nih.gov/33904699/

31. Semwanga AR, Nakubulwa S, Adam T. Applying a system dynamics modelling approach to explore policy options for improving neonatal health in Uganda. Health Res Policy Syst. 2016;14(1):35. Available from: http://health-policy-systems.biomedcentral.com/articles/10.1186/s12961-016-0101-8

32. Shrime MG, Iverson KR, Yorlets R, Roder-Dewan S, Gage AD, Leslie H, et al. Predicted effect of regionalised delivery care on neonatal mortality, utilisation, financial risk, and patient utility in Malawi: an agent-based modelling analysis. Articles Lancet Glob Health. 2019;7:932–71. Available from: https://www.thelancet.com/journals/langlo/article/PIIS2214-109X(19)30170-6/fulltext

33. Pagel C, Lewycka S, Colbourn T, Mwansambo C, Meguid T, Chiudzu G, et al. Estimation of potential effects of improved community-based drug provision, to augment health-facility strengthening, on maternal mortality due to post-partum haemorrhage and sepsis in sub-Saharan Africa: an equity-effectiveness model. Lancet. 2009;374(9699):1441–8. Available from: https://www.thelancet.com/journals/lancet/article/PIIS014067360961566X/fulltext

34. Nadkarni D, Minocha A, Harpaldas H, Kim G, Gopaluni A, Gravelyn S, et al. Predicting resource-dependent maternal health outcomes at a referral hospital in Zanzibar using patient trajectories and mathematical modeling. Brownie SM, editor. PLoS One. 2019;14(3):e0212753. Available from: https://dx.plos.org/10.1371/journal.pone.0212753

35. Michalow J, Chola L, McGee S, Tugendhaft A, Pattinson R, Kerber K, et al. Triple return on investment: the cost and impact of 13 interventions that could prevent stillbirths and save the lives of mothers and babies in South Africa. BMC Pregnancy Childbirth. 2015;15:39. Available from: https://www.ncbi.nlm.nih.gov/pmc/articles/PMC4337184/

36. Memirie ST, Tolla MT, Desalegn D, Hailemariam M, Norheim OF, Verguet S, et al. A cost-effectiveness analysis of maternal and neonatal health interventions in Ethiopia. Health Policy Plan. 2019;34(4):289–97. Available from: https://pubmed.ncbi.nlm.nih.gov/31106346/

37. McPake B, Edoka I, Witter S, Kielmann K, Taegtmeyer M, Dieleman M, et al. Cost-effectiveness of community-based practitioner programmes in Ethiopia, Indonesia and Kenya. Bull World Health Organ. 2015;93(9):631–639A. Available from: https://www.ncbi.nlm.nih.gov/pmc/articles/PMC4581637/

38. McClure EM, Jones B, Rouse DJ, Griffin JB, Kamath-Rayne BD, Downs A, et al. Tranexamic acid to reduce postpartum hemorrhage: A MANDATE systematic review and analyses of impact on maternal mortality. Am J Perinatol. 2015;32(5):469–74.

39. Júnior JM, Cane RM, Gonçalves MP, Sambo J, Konikoff J, Fernandes Q, et al. Projecting the lives saved by continuing the historical scale-up of child and maternal health interventions in Mozambique until 2030. J Glob Health. 2019;9(1):11102. Available from: https://www.ncbi.nlm.nih.gov/pmc/articles/PMC6513506/#:~:text=If%20historical%20trends%20continue%2C%20coverage,constant%20from%202015%20to%202030.

40. LeFevre A, Cabrera-Escobar MA, Mohan D, Eriksen J, Rogers D, Neo Parsons A, et al. Forecasting the Value for Money of Mobile Maternal Health Information Messages on Improving Utilization of Maternal and Child Health Services in Gauteng, South Africa: Cost-Effectiveness Analysis. JMIR Mhealth Uhealth. 2018;6(7):e153. Available from: https://www.ncbi.nlm.nih.gov/pmc/articles/PMC6086931/#:~:text=Conclusions,small%20margins%20of%20coverage%20increases.

41. Kumar MB, Madan JJ, Auguste P, Taegtmeyer M, Otiso L, Ochieng CB, et al. Cost-effectiveness of community health systems strengthening: quality improvement interventions at community level to realise maternal and child health gains in Kenya. BMJ Glob Health. 2021;6(3). Available from: https://gh.bmj.com/content/6/3/e002452

42. Khurmi MS, Sayinzoga F, Berhe A, Bucyana T, Mwali AK, Manzi E, et al. Newborn Survival Case Study in Rwanda - Bottleneck Analysis and Projections in Key Maternal and Child Mortality Rates Using Lives Saved Tool (LiST). Int J MCH AIDS. 2017;6(2):93–108. Available from: https://pubmed.ncbi.nlm.nih.gov/29367886/

43. Kamath-Rayne BD, Griffin JB, Moran K, Jones B, Downs A, McClure EM, et al. Resuscitation and Obstetrical Care to Reduce Intrapartum-Related Neonatal Deaths: A MANDATE Study. Matern Child Health J. 2015;19(8):1853–63. Available from: https://pubmed.ncbi.nlm.nih.gov/25656720/

44. Jo Y, Labrique AB, Lefevre AE, Mehl G, Pfaff T, Walker N, et al. Using the lives saved tool (LiST) to model mHealth impact on neonatal survival in resource-limited settings. PLoS One. 2014;9(7):e102224. Available from: https://journals.plos.org/plosone/article?id=10.1371/journal.pone.0102224

45. Herrick T, Mvundura M, Burke TF, Abu-Haydar E. A low-cost uterine balloon tamponade for management of postpartum hemorrhage: modeling the potential impact on maternal mortality and morbidity in sub-Saharan Africa. BMC Pregnancy Childbirth. 2017 Nov 13;17(1). Available from: https://pubmed.ncbi.nlm.nih.gov/29132342/

46. Griffin JB, McClure EM, Kamath-Rayne BD, Hepler BM, Rouse DJ, Jobe AH, et al. Interventions to reduce neonatal mortality: a mathematical model to evaluate impact of interventions in sub-Saharan Africa. Acta Paediatr. 2017;106(8):1286–95. Available from: https://onlinelibrary.wiley.com/doi/10.1111/apa.13853

47. Griffin JB, Jobe AH, Rouse D, McClure EM, Goldenberg RL, Kamath-Rayne BD. Evaluating WHO-Recommended Interventions for Preterm Birth: A Mathematical Model of the Potential Reduction of Preterm Mortality in Sub-Saharan Africa. Glob Health Sci Pract. 2019;7(2):215–27. Available from: http://www.ghspjournal.org/lookup/doi/10.9745/GHSP-D-18-00402

48. Goldenberg RL, Jones B, Griffin JB, Rouse DJ, Kamath-Rayne BD, Trivedi N, et al. Reducing maternal mortality from preeclampsia and eclampsia in low-resource countries - what should work? Acta Obstet Gynecol Scand. 2015;94(2):148–55. Available from: https://onlinelibrary.wiley.com/doi/10.1111/aogs.12533

49. Goldenberg RL, Griffin JB, Kamath-Rayne BD, Harrison M, Rouse DJ, Moran K, et al. Clinical interventions to reduce stillbirths in sub-Saharan Africa: a mathematical model to estimate the potential reduction of stillbirths associated with specific obstetric conditions. BJOG. 2018;125(2):119–29. Available from: https://pubmed.ncbi.nlm.nih.gov/27704677/

50. Friberg IK, Kinney M V., Lawn JE, Kerber KJ, Odubanjo MO, Bergh AM, et al. Sub-Saharan Africa’s mothers, newborns, and children: how many lives could be saved with targeted health interventions? PLoS Med. 2010;7(6). Available from: https://pubmed.ncbi.nlm.nih.gov/20574515/

51. Epiu I, Alia G, Mukisa J, Tavrow P, Lamorde M, Kuznik A. Estimating the cost and cost-effectiveness for obstetric fistula repair in hospitals in Uganda: a low income country. Health Policy Plan. 2018;33(9):999–1008. Available from: https://www.ncbi.nlm.nih.gov/pmc/articles/PMC6263022/#:~:text=The%20cost%20of%20providing%20fistula,per%20DALY%20averted%20of%20%2454.

52. Curry LA, Byam P, Linnander E, Andersson KM, Abebe Y, Zerihun A, et al. Evaluation of the Ethiopian Millennium Rural Initiative: impact on mortality and cost-effectiveness. PLoS One. 2013;8(11):e79847. Available from: https://pubmed.ncbi.nlm.nih.gov/24260307/

53. Chola L, Pillay Y, Barron P, Tugendhaft A, Kerber K, Hofman K. Cost and impact of scaling up interventions to save lives of mothers and children: taking South Africa closer to MDGs 4 and 5. Glob Health Action. 2015;8(1). Available from: https://pubmed.ncbi.nlm.nih.gov/25906769/

54. Chola L, Fadnes LT, Engebretsen IMS, Nkonki L, Nankabirwa V, Sommerfelt H, et al. Cost-Effectiveness of Peer Counselling for the Promotion of Exclusive Breastfeeding in Uganda. PLoS One. 2015;10(11). Available from: https://journals.plos.org/plosone/article?id=10.1371/journal.pone.0142718

55. Carvalho N, Hoque ME, Oliver VL, Byrne A, Kermode M, Lambert P, et al. Cost-effectiveness of inhaled oxytocin for prevention of postpartum haemorrhage: a modelling study applied to two high burden settings. BMC Med. 2020;18(1):201. Available from: https://bmcmedicine.biomedcentral.com/articles/10.1186/s12916-020-01658-y

56. Bhutta ZA, Das JK, Rizvi A, Gaffey MF, Walker N, Horton S, et al. Evidence-based interventions for improvement of maternal and child nutrition: What can be done and at what cost? The Lancet. 2013;382(9890):452–77. Available from: http://www.thelancet.com/article/S0140673613609964/fulltext

57. Bartlett L, Weissman E, Gubin R, Patton-Molitors R, Friberg IK. The impact and cost of scaling up midwifery and obstetrics in 58 low- and middle-income countries. PLOS One. 2014;9(6):e98550. Available from: https://journals.plos.org/plosone/article?id=10.1371/journal.pone.0098550

58. Ali A, Nudel J, Heberle CR, Santorino D, Olson KR, Hur C. Cost effectiveness of a novel device for improving resuscitation of apneic newborns. BMC Pediatr. 2020;20(1):46. Available from: https://bmcpediatr.biomedcentral.com/articles/10.1186/s12887-020-1925-5

59. David S, Abbas-Chorfa F, Vanhems P, Vallin B, Iwaz J, Ecochard R. Promotion of WHO feeding recommendations: a model evaluating the effects on HIV-free survival in African children. J Hum Lact. 2008;24(2):140–9. Available from: https://pubmed.ncbi.nlm.nih.gov/18436965/

60. Darmstadt GL, Walker N, Lawn JE, Bhutta ZA, Haws RA, Cousens S. Saving newborn lives in Asia and Africa: cost and impact of phased scale-up of interventions within the continuum of care. Health Policy Plan. 2008;23(2):101–17. Available from: https://academic.oup.com/heapol/article/23/2/101/592767

61. Bradley SEK, Prata N, Young-Lin N, Bishai DM. Cost-effectiveness of misoprostol to control postpartum hemorrhage in low-resource settings. International Journal of Gynecology & Obstetrics. 2007;97(1):52–6. Available from: http://doi.wiley.com/10.1016/j.ijgo.2006.12.005

62. Baggaley RF, Burgin J, Campbell OMR. The potential of medical abortion to reduce maternal mortality in Africa: what benefits for Tanzania and Ethiopia? PLoS One. 2010;5(10):e13260. Available from: https://journals.plos.org/plosone/article?id=10.1371/journal.pone.0013260

63. Chou VB, Walker N, Kanyangarara M. Estimating the global impact of poor quality of care on maternal and neonatal outcomes in 81 low- and middle-income countries: A modeling study. PLoS Med. 2019;16(12):e1002990. Available from: https://pubmed.ncbi.nlm.nih.gov/31851685/

64. Nove A, Friberg IK, de Bernis L, McConville F, Moran AC, Najjemba M, et al. Potential impact of midwives in preventing and reducing maternal and neonatal mortality and stillbirths: a Lives Saved Tool modelling study. Lancet Glob Health. 2021;9(1):e24–32. Available from: https://www.thelancet.com/journals/langlo/article/PIIS2214-109X(20)30397-1/fulltext

65. Harrison MS, Griffin JB, McClure EM, Jones B, Moran K, Goldenberg RL. Maternal Mortality from Obstructed Labor: A MANDATE Analysis of the Ability of Technology to Save Lives in Sub-Saharan Africa. Am J Perinatol. 2016;33(9):873–81. Available from: https://pubmed.ncbi.nlm.nih.gov/27031054/

66. Hitimana R, Lindholm L, Mogren I, Krantz G, Nzayirambaho M, Sengoma JPS, et al. Incremental cost and health gains of the 2016 WHO antenatal care recommendations for Rwanda: results from expert elicitation. Health Res Policy Syst. 2019;17(1):36. Available from: https://health-policy-systems.biomedcentral.com/articles/10.1186/s12961-019-0439-9

67. Fotso JC, Ambrose A, Hutchinson P, Ali D. Improving maternal and newborn care: cost-effectiveness of an innovation to rebrand traditional birth attendants in Sierra Leone. Int J Public Health. 2020;65(9):1603–12. Available from: https://pubmed.ncbi.nlm.nih.gov/33037894/

68. Erim DO, Resch SC, Goldie SJ. Assessing health and economic outcomes of interventions to reduce pregnancy-related mortality in Nigeria. BMC Public Health. 2012 Sep 14;12(1):1–11. Available from: https://bmcpublichealth.biomedcentral.com/articles/10.1186/1471-2458-12-786

69. McGee SA, Chola L, Tugendhaft A, Mubaiwa V, Moran N, McKerrow N, et al. Strategic planning for saving the lives of mothers, newborns and children and preventing stillbirths in KwaZulu-Natal province South Africa: modelling using the Lives Saved Tool (LiST). BMC Public Health. 2016;16:49. Available from: https://bmcpublichealth.biomedcentral.com/articles/10.1186/s12889-015-2661-x

70. Keita Y, Sangho H, Roberton T, Vignola E, Traoré M, Munos M. Using the Lives Saved Tool to aid country planning in meeting mortality targets: a case study from Mali. BMC Public Health. 2017;17(Suppl 4):777. Available from: https://bmcpublichealth.biomedcentral.com/articles/10.1186/s12889-017-4749-y

71. Chou VB, Friberg IK, Christian M, Walker N, Perry HB. Expanding the population coverage of evidence-based interventions with community health workers to save the lives of mothers and children: an analysis of potential global impact using the Lives Saved Tool (LiST). J Glob Health. 2017;7(2):20401. Available from: https://pubmed.ncbi.nlm.nih.gov/28959436/

72. Homer CSE, Friberg IK, Dias MAB, ten Hoope-Bender P, Sandall J, Speciale AM, et al. The projected effect of scaling up midwifery. Lancet. 2014;384(9948):1146–57. Available from: https://www.thelancet.com/journals/lancet/article/PIIS0140-6736(14)60790-X/fulltext

73. Bhutta ZA, Yakoob MY, Lawn JE, Rizvi A, Friberg IK, Weissman E, et al. Stillbirths: what difference can we make and at what cost? Lancet. 2011;377(9776):1523–38. Available from: https://pubmed.ncbi.nlm.nih.gov/21496906/

74. Marsh A, Munos M, Baya B, Sanon D, Gilroy K, Bryce J. Using LiST to model potential reduction in under-five mortality in Burkina Faso. BMC Public Health. 2013;13 Suppl 3(Suppl 3). Available from: https://pubmed.ncbi.nlm.nih.gov/24564341/

75. Ruhago GM, Ngalesoni FN, Norheim OF. Addressing inequity to achieve the maternal and child health millennium development goals: looking beyond averages. BMC Public Health. 2012;12:1119. Available from: https://bmcpublichealth.biomedcentral.com/articles/10.1186/1471-2458-12-1119

76. Onarheim KH, Tessema S, Johansson KA, Eide KT, Norheim OF, Miljeteig I. Prioritizing child health interventions in Ethiopia: modeling impact on child mortality, life expectancy and inequality in age at death. PLoS One. 2012;7(8). Available from: https://pubmed.ncbi.nlm.nih.gov/22879890/

77. Chou VB, Bubb-Humfryes O, Sanders R, Walker N, Stover J, Cochrane T, et al. Pushing the envelope through the Global Financing Facility: potential impact of mobilising additional support to scale-up life-saving interventions for women, children and adolescents in 50 high-burden countries. BMJ Glob Health. 2018;3(5):e001126. Available from: https://gh.bmj.com/content/3/5/e001126

78. Walker N, Yenokyan G, Friberg IK, Bryce J. Patterns in coverage of maternal, newborn, and child health interventions: projections of neonatal and under-5 mortality to 2035. Lancet. 2013;382(9897):1029–38. Available from: https://pubmed.ncbi.nlm.nih.gov/24054534/

79. Ruducha J, Mann C, Singh NS, Gemebo TD, Tessema NS, Baschieri A, et al. How Ethiopia achieved Millennium Development Goal 4 through multisectoral interventions: a Countdown to 2015 case study. Lancet Glob Health. 2017;5(11):e1142–51. Available from: https://www.thelancet.com/journals/langlo/article/PIIS2214-109X(17)30331-5/fulltext

80. Keats EC, Ngugi A, Macharia W, Akseer N, Khaemba EN, Bhatti Z, et al. Progress and priorities for reproductive, maternal, newborn, and child health in Kenya: a Countdown to 2015 country case study. Lancet Glob Health. 2017;5(8):e782–95. Available from: https://pubmed.ncbi.nlm.nih.gov/28716350/

81. Afnan-Holmes H, Magoma M, John T, Levira F, Msemo G, Armstrong CE, et al. Tanzania’s countdown to 2015: an analysis of two decades of progress and gaps for reproductive, maternal, newborn, and child health, to inform priorities for post-2015. Lancet Glob Health. 2015;3(7):e396–409. Available from: https://www.thelancet.com/journals/langlo/article/PIIS2214-109X(15)00059-5/fulltext

82. Macicame I, Magaço A, Cassocera M, Amado C, Feriano A, Chicumbe S, et al. Intervention heroes of Mozambique from 1997 to 2015: estimates of maternal and child lives saved using the Lives Saved Tool. J Glob Health. 2018;8(2):21202. Available from: https://www.ncbi.nlm.nih.gov/pmc/articles/PMC6300161/

83. Niyeha D, Malamsha D, Mpembeni R, Charwe D, Epimark S, Malima K, et al. Explaining progress towards Millennium Development Goal 4 for child survival in Tanzania. J Glob Health. 2018;8(2):21201. Available from: https://www.ncbi.nlm.nih.gov/pmc/articles/PMC6319734/

84. Amouzou A, Habi O, Bensaïd K. Reduction in child mortality in Niger: a Countdown to 2015 country case study. Lancet. 2012;380(9848):1169–78. Available from: https://pubmed.ncbi.nlm.nih.gov/22999428/#:~:text=Findings%3A%20The%20mortality%20rate%20in,decline%20of%205%C2%B71%25.

85. Kanyuka M, Ndawala J, Mleme T, Chisesa L, Makwemba M, Amouzou A, et al. Malawi and Millennium Development Goal 4: a Countdown to 2015 country case study. Lancet Glob Health. 2016;4(3):e201–14. Available from: https://pubmed.ncbi.nlm.nih.gov/26805586/

86. Ward ZJ, Atun R, King G, Sequeira Dmello B, Goldie SJ. A simulation-based comparative effectiveness analysis of policies to improve global maternal health outcomes. Nat Med. 2023;29(5):1262–72. Available from: 10.1038/s41591-023-02310-x

87. Petrou S, Gray A. Economic evaluation using decision analytical modelling: design, conduct, analysis, and reporting. BMJ. 2011;342(7808). Available from: https://www.bmj.com/content/342/bmj.d1766

88. Rautenberg T, Gerritsen A, Downes M. Health Economic Decision Tree Models of Diagnostics for Dummies: A Pictorial Primer. Diagnostics (Basel). 2020;10(3). Available from: https://pubmed.ncbi.nlm.nih.gov/32183372/

89. Johns Hopkins Bloomberg School of Public Health. LiST Visualizer. 2023. Available from: https://listvisualizer.org/

90. Research Triangle Institute International. MANDATE: Maternal and Neonatal Directed Assessment of Technology. 2014. MANDATE: Condition Maps. Available from: http://www.mandate4mnh.org/help/maps

91. Stegmuller AR, Self A, Litvin K, Roberton T. How is the Lives Saved Tool (LiST) used in the global health community? Results of a mixed-methods LiST user study. BMC Public Health. 2017;17(4):143–50. Available from: https://bmcpublichealth.biomedcentral.com/articles/10.1186/s12889-017-4750-5

92. Walker N, Tam Y, Friberg IK. Overview of the Lives Saved Tool (LiST). BMC Public Health. 2013;13(Suppl 3):S1. Available from: http://www.jhsph.edu/IIP/list

93. Sturmberg J, Lanham HJ. Understanding health care delivery as a complex system: achieving best possible health outcomes for individuals and communities by focusing on interdependencies. J Eval Clin Pract. 2014;20(6):1005–9. Available from: https://pubmed.ncbi.nlm.nih.gov/24797788/

94. Lipsitz LA, Seniorlife H, Israel B. Understanding Health Care as a Complex System: The Foundation for Unintended Consequences. JAMA. 2012;308(3):243–4. Available from: https://www.ncbi.nlm.nih.gov/pmc/articles/PMC3511782/

95. Martínez-García M, Hernández-Lemus E. Health Systems as Complex Systems. American Journal of Operations Research. 2013;2013(01):113–26. Available from: https://www.scirp.org/journal/paperinformation.aspx?paperid=27538

96. Fink G, Cohen J. Delivering quality: safe childbirth requires more than facilities. Lancet Glob Health. 2019.;7(8):e990–1. Available from: https://www.thelancet.com/journals/langlo/article/PIIS2214-109X(19)30193-7/fulltext#:~:text=Facility%20birth%20does%20not%20necessarily,of%20safe%2Dguarding%20uncomplicated%20births.

97. Gabrysch S, Nesbitt RC, Schoeps A, Hurt L, Soremekun S, Edmond K, et al. Does facility birth reduce maternal and perinatal mortality in Brong Ahafo, Ghana? A secondary analysis using data on 119 244 pregnancies from two cluster-randomised controlled trials. Lancet Glob Health. 2019.;7(8):e1074–87. Available from: https://www.thelancet.com/journals/langlo/article/PIIS2214-109X(19)30165-2/fulltext

98. de Savigny D & Adam T. Systems thinking for health systems strengthening. 2009. Available from: https://apps.who.int/iris/handle/10665/44204

99. Paina L, Peters DH. Understanding pathways for scaling up health services through the lens of complex adaptive systems. Health Policy Plan. 2012 Aug;27(5):365–73. Available from: https://pubmed.ncbi.nlm.nih.gov/21821667

100. Verguet S, Feldhaus I, Jiang Kwete X, Aqil A, Atun R, Bishai D, et al. Health system modelling research: towards a whole-health-system perspective for identifying good value for money investments in health system strengthening. BMJ Glob Health. 2019;4(2):e001311. Available from: https://gh.bmj.com/content/4/2/e001311

101. Briggs ADM, Wolstenholme J, Blakely T, Scarborough P. Choosing an epidemiological model structure for the economic evaluation of non-communicable disease public health interventions. Popul Health Metr. 2016;14(1):1–12. Available from: https://pophealthmetrics.biomedcentral.com/articles/10.1186/s12963-016-0085-1

102. Tracy M, Cerdá M, Keyes KM. Agent-Based Modeling in Public Health: Current Applications and Future Directions. Annu Rev Public Health. 2018;39:77–94. Available from: https://pubmed.ncbi.nlm.nih.gov/29328870/

103. Willem L, Verelst F, Bilcke J, Hens N, Beutels P. Lessons from a decade of individual-based models for infectious disease transmission: A systematic review (2006-2015). BMC Infect Dis. 2017;17(1):1–16. Available from: https://bmcinfectdis.biomedcentral.com/articles/10.1186/s12879-017-2699-8

104. Marshall BDL. Agent-Based Modeling. In: El-Sayed A, Galea S (ed.) Systems Science and Population Health. Oxford University Press; 2017. p. 87–98. Available from: https://oxford.universitypressscholarship.com/view/10.1093/acprof:oso/978019 0492397.001.0001/acprof-9780190492397-chapter-8

105. Marshall BDL. Agent-Based Modelling. In: Lash T, VanderWeele T, Haneuse S, Rothman K, editors. Modern Epidemiology - Fourth Edition. Philideliphia: Wolters Kluwer Publications; 2021. p. 785.

106. Gilbert N. The Idea of Agent-Based Modeling. In: Agent-Based Models. California: SAGE Publications, Inc.; 2020. p. 1–23. Available from: https://methods.sagepub.com/book/agent-based-models-2e/i214.xml

107. Hammond RA. Overview of Current Concepts and Process for Agent-Based Modeling. In: New Horizons in Modeling and Simulation for Social Epidemiology and Public Health. John Wiley & Sons, Ltd; 2021. p. 31–43. Available from: https://onlinelibrary.wiley.com/doi/full/10.1002/9781118589397.ch3

108. Shoveller J, Viehbeck S, Di Ruggiero E, Greyson D, Thomson K, Knight R. A critical examination of representations of context within research on population health interventions. Crit. Public Health 2015;26(5):487–500. Available from: https://www.tandfonline.com/doi/abs/10.1080/09581596.2015.1117577?journalCode=ccph20

109. Adams S, Rhodes T, Lancaster K. New directions for participatory modelling in health: Redistributing expertise in relation to localised matters of concern. Glob Public Health. 2021;17(9):1827–1841. Available from: https://www.tandfonline.com/doi/citedby/10.1080/17441692.2021.1998575?scroll=top&needAccess=true&role=tab

110. Baugh Littlejohns L, Baum F, Lawless A, Freeman T. The value of a causal loop diagram in exploring the complex interplay of factors that influence health promotion in a multisectoral health system in Australia. Health Res Policy Syst. 2018;16(1):1–12. Available from: https://health-policy-systems.biomedcentral.com/articles/10.1186/s12961-018-0394-x#:~:text=Creating%20a%20causal%20loop%20diagram,and%20practice%20can%20be%20improved.

111. Sargent RG. Verification and validation of simulation models. Journal of Simulation. 2013;7(1):12–24. Available from: https://www.tandfonline.com/action/journalInformation?journalCode=tjsm20

112. World Health Organization. WHO recommendations: Intrapartum care for a positive childbirth experience.. 2018 Feb. Available from: https://www.who.int/publications/i/item/9789241550215

113. World Health Organization. WHO recommendations on antenatal care for a positive pregnancy experience. 2016. Available from: WHO recommendations on antenatal care for a positive pregnancy experience

114. World Health Organization. WHO recommendations on maternal and newborn care for a positive postnatal experience. 2022. Available from: https://www.who.int/publications/i/item/9789240045989

115. Lawn JE, Blencowe H, Pattinson R, Cousens S, Kumar R, Ibiebele I, et al. Stillbirths: Where? When? Why? How to make the data count? Lancet. 2011;377(9775):1448–63. Available from: https://pubmed.ncbi.nlm.nih.gov/21496911/

116. Frøen JF, Cacciatore J, McClure EM, Kuti O, Jokhio AH, Islam M, et al. Stillbirths: Why they matter. Lancet. 2011;377(9774):1353–66. Available from: https://pubmed.ncbi.nlm.nih.gov/21496911/

117. Qureshi ZU, Millum J, Blencowe H, Kelley M, Fottrell E, Lawn JE, et al. Stillbirth should be given greater priority on the global health agenda. BMJ. 2015;351. Available from: https://www.bmj.com/content/351/bmj.h4620

118. Frøen JF, Friberg IK, Lawn JE, Bhutta ZA, Pattinson RC, Allanson ER, et al. Stillbirths: Progress and unfinished business. Lancet. 2016; 387(10018):574–86. Available from: https://www.thelancet.com/journals/lancet/article/PIIS0140-6736(15)00818-1/fulltext

119. Westby CL, Erlandsen AR, Nilsen SA, Visted E, Thimm JC. Depression, anxiety, PTSD, and OCD after stillbirth: a systematic review. BMC Pregnancy Childbirth. 2021 ;21(1). Available from: /pmc/articles/PMC8600867/

120. Ota E, da Silva Lopes K, Middleton P, Flenady V, Wariki WMV, Rahman MO, et al. Antenatal interventions for preventing stillbirth, fetal loss and perinatal death: an overview of Cochrane systematic reviews. Cochrane Database Syst Rev. 2020;2020(12). Available from: https://www.cochranelibrary.com/cdsr/doi/10.1002/14651858.CD009599.pub2/full

121. Wastnedge E, Waters D, Murray SR, McGowan B, Chipeta E, Nyondo-Mipando AL, et al. Interventions to reduce preterm birth and stillbirth, and improve outcomes for babies born preterm in low- and middle-income countries: A systematic review. J Glob Health. 2021;11. Available from: https://pubmed.ncbi.nlm.nih.gov/35003711/

122. Paez A. Grey literature: An important resource in systematic reviews. J Evid Based Med.2017;10(3):233–240. Available from: https://pubmed.ncbi.nlm.nih.gov/28857505/

123. Walker N, Friberg IK. Introduction: reporting on updates in the scientific basis for the Lives Saved Tool (LiST). BMC public health. 2017;17(Suppl 4):774. Available from: https://pubmed.ncbi.nlm.nih.gov/29143617/

124. Godin K, Stapleton J, Kirkpatrick SI, Hanning RM, Leatherdale ST. Applying systematic review search methods to the grey literature: A case study examining guidelines for school-based breakfast programs in Canada. Syst Rev. 2015;4(1):1–10. Available from: https://pubmed.ncbi.nlm.nih.gov/26494010/

125. Abuelezam NN, Rough K, Seage GR. Individual-Based Simulation Models of HIV Transmission: Reporting Quality and Recommendations. PLoS One. 2013;8(9). Available from: https://journals.plos.org/plosone/article?id=10.1371/journal.pone.0075624

126. Giddings R, Indravudh P, Medley GF, Bozzani F, Gafos M, Malhotra S, et al. Infectious Disease Modelling of HIV Prevention Interventions: A Systematic Review and Narrative Synthesis of Compartmental Models. Pharmacoeconomics. 2023;41(6):693–707. Available from: https://link.springer.com/article/10.1007/s40273-023-01260-z

127. Colquhoun HL, Levac D, O’Brien KK, Straus S, Tricco AC, Perrier L, et al. Scoping reviews: time for clarity in definition, methods, and reporting. J Clin Epidemiol. 2014;67(12):1291–4. Available from: https://pubmed.ncbi.nlm.nih.gov/25034198/

